# Q Fever Related Community Infections, US Exposure to *Coxiella burnetii*

**DOI:** 10.1101/2025.03.23.25324484

**Authors:** Charles F. Dillon, Gwendolyn R. Dillon

## Abstract

*Coxiella burnetii* is a significant infectious pathogen that causes Q fever. Q fever is thought to be uncommon in the US and most human cases are believed to occur in agricultural livestock workers. However, the extent of community exposure to *C. burnetii* isn’t known with certainty. Using nationally representative US National Health and Nutrition Examination Survey serologic, demographic and occupational history data, the magnitude of US adult general population exposure to *C. burnetii* excluding agricultural sector workers was estimated. Exposure was defined as positive serum IgG antibodies by immunofluorescence assay (e.g. current or past infection). 3.0% (95% CI 2.0-4.4) of the US population met criteria for *C. burnetii* exposure, some 6.2 million persons. Overall, 86.9% (95%CI: 75.5-98.4) of seropositives had no lifetime history of work in the agricultural sector (5.5 million persons). This was consistently true across all US demographic groups: age 20-59 years 87.3%, age 60+ years 85.7%, men 86.1%, women 87.6%, Non-Hispanic Whites 82%, Non-Hispanic Blacks 95.8%, Mexican Americans 89.4%, immigrants from Mexico 83.5% and other immigrants 96.8%. As half of *C. burnetii* infections result in acute Q fever and chronic Q fever conveys significant mortality, community level risks to the general public may be significant.

## 1. Introduction

*Coxiella burnetii* is an important pathogen, the causal agent of Q fever, one of the select US national notifiable infectious diseases and a significant but treatable health hazard [1-3]. It is a potent gram-negative intracellular pathogen with a low infectious dose [4,5]. Half of those infected develop clinical illness, typically an acute respiratory illness or hepatitis. Infection can also be followed by chronic illness which carries significant mortality risk [1,6]. *C. burnetii* is hardy and capable of persisting in the environment for months to years, significantly increasing population exposure risks [7]. A primary route of transmission to humans is airborne inhalation of bacteria from infected animal birth products or contaminated dust and soils.^1^

A major reservoir for *C. burnetii* is farm animals, specifically domesticated ruminants including cattle, sheep and goats [8,9]. However, since livestock workers are a small minority of the general population, Q fever has usually been considered uncommon in the US, a rare illness mainly affecting this one specific occupational group. The fact that relatively few Q fever cases are officially reported each year to the US National Notifiable Diseases Surveillance System (NNDSS) may have served to strengthen this perception (194 cases in 2022) [2]. However, Q fever in the US is considered a largely unreported disease [1,8,10].

How accurate is the impression that *C. burnetii* infection and clinical disease are mainly restricted to the agricultural industry? A review of Q fever case reports submitted to the NNDSS from 2000 to 2010 show that 79% of officially notified Q fever cases had no history of work in traditional high-risk occupations and 60% reported no contact with livestock [1]. These estimates could be a signal of significant *C. burnetii* exposures occurring in the US general population, however the reports were based on a voluntary reporting system (passive surveillance) that was not nationally representative.

There is nationally representative data for the US prevalence of positive *C. burnetii* antibodies. A US National Health and Nutrition Examination Survey (NHANES) surplus sera study using 2003-04 specimens showed that some 3.1% of the US adult population were *C. burnetii* seropositive, an estimated 6.1 million persons [11]. These numbers are remarkable, indicating that exposure to and infection with *C. burnetii* is in fact common in the US. It also suggests that the prevalence of US Q fever cases could be much greater than officially notified case reports. Further, NHANES routinely obtains nationally representative work history data [12]. This captures the greater part of a participant’s lifetime employment. However, the 2-year survey dataset was the minimum sample size for any NHANES analysis. Given an overall 3% *C. burnetii* seropositive rate, stable statistical estimates for occupational exposure analyses for many specific occupations including agricultural workers were not possible.

It was not appreciated, however, that the statistical complement of the *C. burnetii* infection rate in agricultural workers, the seroprevalence in persons with no prior history of work in the agricultural sector, can be reliably estimated from the NHANES data and with precision. This seroprevalence estimate is presented here for the US and its major demographic subgroups. This metric is important as it can potentially provide an initial perspective on the scope of US population-level exposures to the general public outside the agricultural sector.

## 2. Materials and Methods

NHANES is a series of cross-sectional, active surveillance surveys monitoring the health and nutritional status of the ambulatory, noninstitutionalized US civilian population. Each 2-year survey cycle is nationally representative. Data is collected by in-person household interviews with examinations and laboratory studies performed in mobile examination centers [13]. NHANES is a demographically based survey that uses a complex, multistage, survey design to estimate national level prevalences [14]. Oversampling is used to capture adequate data for key demographic sub-groups. The NHANES 2003-04 survey had a 70% adult health examination response rate [15].

### 2.1 NHANES Occupational data

NHANES fielded one of the standardized US National Center for Health Statistics (NCHS) occupational history surveillance instruments [12]. Data was collected by trained professional interviewers with in-field data quality control. Data collection also included demographic data (age, sex, ethnicity, and nativity). Data for the participant’s current and longest held jobs was also collected, and if applicable, data for an individual’s job during which their asthma had begun if not the current or longest held position. This captures the greater part of an individual’s work history but is not a complete listing of all jobs ever held. The NHANES dataset did not have a specific variable for work with livestock. The NHANES industry and occupation text data were coded by NCHS Division of Vital Statistics staff using the U.S. Census Bureau’s 2000 version of its Occupation and Industry Coding System [16-17]. For public release, the detailed US Census data codes were abstracted into 45 industry and 41 occupation groups [16] (Appendix A). A participant was classified as having worked in the agricultural sector if there was an industry or occupation code to indicate it. Industry codes included agricultural production, support services and forestry; occupation codes included farm operators, managers, supervisors, farm and nursery workers and related agricultural occupations [16] (Appendix A).

Statistical analysis for most individual occupational titles was not possible due to small sample sizes, even when the data was regrouped into 17 occupational categories (Supplemental Table 1). We therefore condensed the data into four major socioeconomic groups using an occupational group classification system previously used by the US Centers for Disease Control and Prevention [18] (p. 231). These were professional, technical and office workers (occupation group codes 1-16); service workers (codes 17-24); agriculture and related work (codes 25-27); and factory, repair, construction, transport, freight/materials work [16] (codes 28-40). We also added categories for those who reported a history of work in more than one of the occupational groups and for those who had never worked. To assess whether our occupational data had sufficient sample size for the analyses, the total study person-years of work history were computed. For quality assurance purposes, age-specific NHANES 2003-04 civilian labor force participation rates were compared to known US Bureau of Labor Statistics (BLS) values [19], Supplemental Tables 2,3).

### 2.2 C. burnetii serology data

The 2022 update of the NHANES 2003-04 *C. burnetii* stored surplus sera data was used for analysis [20]. Sera were screened for *C. burnetii* IgG antibodies (N=4,236) using an enzyme-linked immunosorbent assay (ELISA). Positive or equivocal ELISA results were then tested for IgG Phase 1 and 2 antibodies by immunofluorescence assay using the Philip et al. method adapted to *C. burnetii* [21-22] (purified Phase 1 Phase 2, strain Nine Mile; Rocky Mountain Laboratories). Exposure in this study was defined as a positive Phase 1 or 2 serum IgG antibody titer of ≥ 1:16 (e.g. current or past infection).

### 2.3 Statistical methods

Data assembly and statistical analysis utilized SAS^™^ (release 9.4, SAS Institute, Inc., Cary, NC) and SUDAAN^™^ (release 11.0.1, Research Triangle Institute, NC). Survey design variables (strata and primary sampling units) and health examination sample weights were used to account for differential probabilities of participant selection, to adjust for survey nonresponse and noncoverage, and to provide nationally representative estimates. Standard errors were estimated using Taylor series linearization. Prevalence estimates were age-adjusted using direct standardization. T-tests were used to compare estimates with p ≤.05 considered significant. NCHS criteria and software were employed to assess the statistical reliability of estimated prevalences and proportions, based on effective sample size, relative confidence interval widths and degrees of freedom [23-25]. Where statistical reliability criteria were not met, select prevalence estimates without confidence intervals are presented for perspective.

## 3. Results

### 3.1 Overall US C. burnetii seroprevalence

Overall, 175 of 4,236 survey participants were *C. burnetii* seropositive, an estimated 3.0% US prevalence (95%CI 2.0-4.4) (Table 1). This corresponds to 6.2 million adults in the general popu^-^ lation (95%CI 4.1-9.0 million). Infection risk was significantly increased at older ages: 4.2% in those ages 60+ years vs. 2.7% in younger adults (t=2.52, p=.02). Men showed higher prevalence of infection than women 3.8% vs. 2.3% (t=2.23, p=.04), however the numbers of individuals affected in both sexes was substantial, 3.8 million infections among men (95%CI 2.5-5.5) and 2.5 million among women (95%CI 1.4-4.1). Mexican Americans had significantly increased risk of infection, 7.5% compared to 2.8% in Non-Hispanic Whites (t=5.71, p<.01). Seroprevalence was 1.4% in the Non-Hispanic Black population (95%CI 0.6-2.8). 2.4% (95% CI 1.4-3.6) of US born participants had positive antibodies whereas foreign born participants had higher infection rates: 8.6% (95%CI 6.2.-11.5) of those born in Mexico and 6.9% (95%CI 3.6-11.7) of those born in other countries were positive.

**Table 1.**
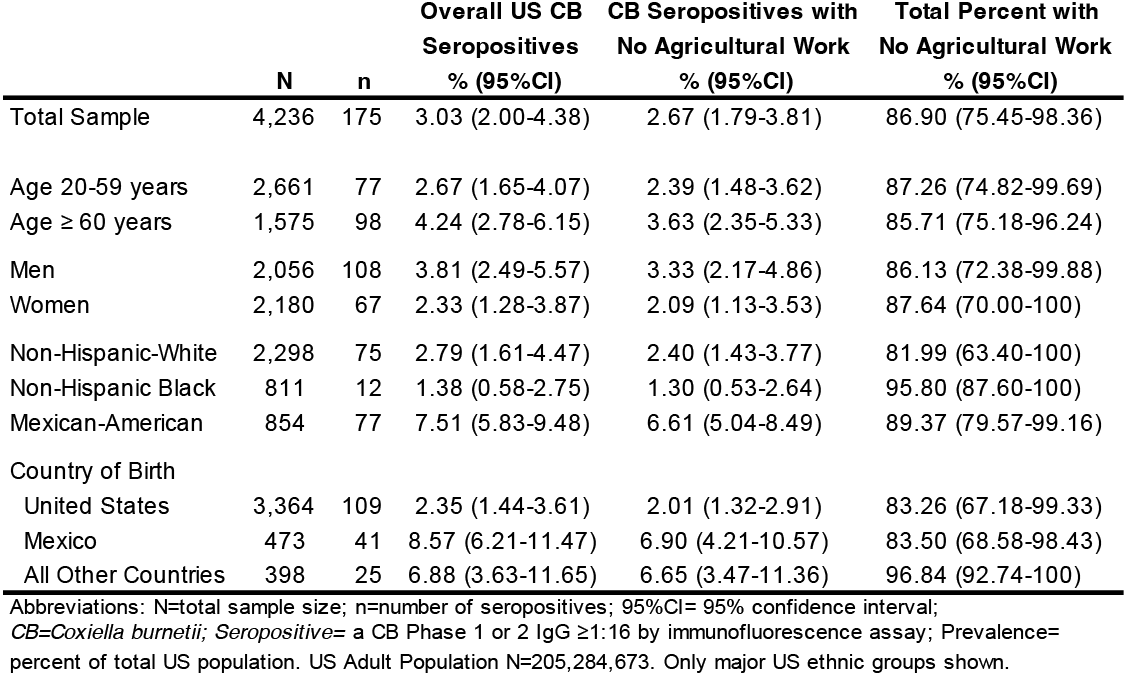
*Coxiella burnetii* Seroprevalence in US Adults with No History of Agricultural Work.

### 3.2 Occupational sample data descriptives

NHANES 2003-04 dataset had a total of 59,214 person-years of work for the occupational history analysis, 1 person-year defined as one year an individual worked (Table 2). There were a total of 25,410 person-years of work among those who were currently working, 732 person-years among the unemployed and 32,072 person-years worked for those who were not currently working. The latter included those retired (24,022 person-years), the disabled, homemakers, personal illness, and others (Supplemental Table 4).

**Table 2.** NHANES 2003-04 Overall and Age-Specific Person-Years of Work Counts.

|  | Total PYR | Currently Employed | Unemployed | Not Working |
| --- | --- | --- | --- | --- |
| **Total All Ages:** | 59,214 | 25,410 | 732 | 33,072 |
| 20-24 yrs | 966 | 717 | 41 | 208 |
| 25-34 yrs | 3,750 | 2,930 | 156 | 664 |
| 35-44 yrs | 6,372 | 5,043 | 157 | 1,172 |
| 45-54 yrs | 9,092 | 6,924 | 152 | 2,016 |
| 55-64 yrs | 10,601 | 5,304 | 71 | 5,226 |
| 65 + yrs | 28,433 | 4,492 | 155 | 23,786 |

The NHANES 2003-04 *Coxiella burnetii* serology subsample is US adults ages 20+ years.
Abbreviations: PYR= person-years; 1 person-year= one year an individual has worked.

The overall NHANES 2003-04 US civilian labor force participation rate in the study sample was 66.5% (95%CI 64.3-68.7) (Table 3). This was consistent with the Bureau of Labor Statistics estimates for 2003 (66.2%) and 2004 (66.0%). The NHANES age-specific labor force participation rates for the current study sample age range (adults 20+ years) were also consistent with BLS estimates. As BLS reports do not routinely provide standard error estimates, the NHANES and BLS age-specific estimates were not further compared.

**Table 3.** US Labor Force Participation Rates: BLS Compared to NHANES 2003-04.

|  | BLS 2003<br>Estimates | BLS 2004<br>Estimates | 2003-04<br>Sample<br>N | NHANES 2003-04<br>Estimates<br>% (95%CI) |
| --- | --- | --- | --- | --- |
| **Age-Specific Rates** | **%** | **%** |  |  |
| Overall 16+ Years | 66.2 | 66.0 | 5,661 | 66.5 (64.3-68.7) |
| 16-19 yrs | 44.5 | 43.9 | 919 | 53.8 (45.3-62.0) |
| 20-24 yrs | 75.4 | 75.0 | 458 | 72.4 (64.7-78.9) |
| 25-34 yrs | 82.9 | 82.7 | 845 | 78.3 (76.1-80.3) |
| 35-44 yrs | 83.9 | 83.6 | 756 | 80.6 (77.1-83.7) |
| 45-54 yrs | 82.1 | 81.8 | 702 | 75.9 (71.2-80.2) |
| 55-64 yrs | 62.4 | 62.3 | 609 | 56.4 (50.9-61.9) |
| 65+ yrs | 14.0 | 14.4 | 1,372 | 16.6 (13.9-19.8) |

BLS= US Bureau of Labor Statistics; 95%CI= 95% confidence interval; Labor Force Participation Rate is the sum of those currently working or actively working for work divided by the total US civilian non-institutional population. Standard errors not published in BLS reports.

### 3.3 C. burnetii seroprevalence in those with no history of agricultural work

Table 1 and Figure 1 also show the population prevalences and percentages of *C. burnetii* seropositive participants who had no previous history of agricultural work. The US adult population *C. burnetii* seroprevalence in the group with no history of agricultural work was 2.7% (95%CI 1.8-3.8). This was 86.9% (95%CI 75.5-98.4) of all study *C. burnetii* seropositives, equivalent to 5.5 million persons (95%CI 3.7-7.8). Results for the detailed demographic sub-groups were similar. 87.3% of seropositive US adults ages 20-59 years, and 85.7% of adults ages 60+ years reported no history of work in agriculture. Also, seropositive men and women had similar rates for no history of agricultural work: 86.1% (95% CI 72.4-99.9) for men and 87.6% for women (95%CI 70.0-100). As a group, 89.4% (95%CI 79.6-99.2) of seropositive Mexican Americans reported no history of prior agricultural work. Further, 83.5% (95% CI 68.6-98.4) of seropositive persons born in Mexico reported no history of agricultural work.

**Figure 1.**
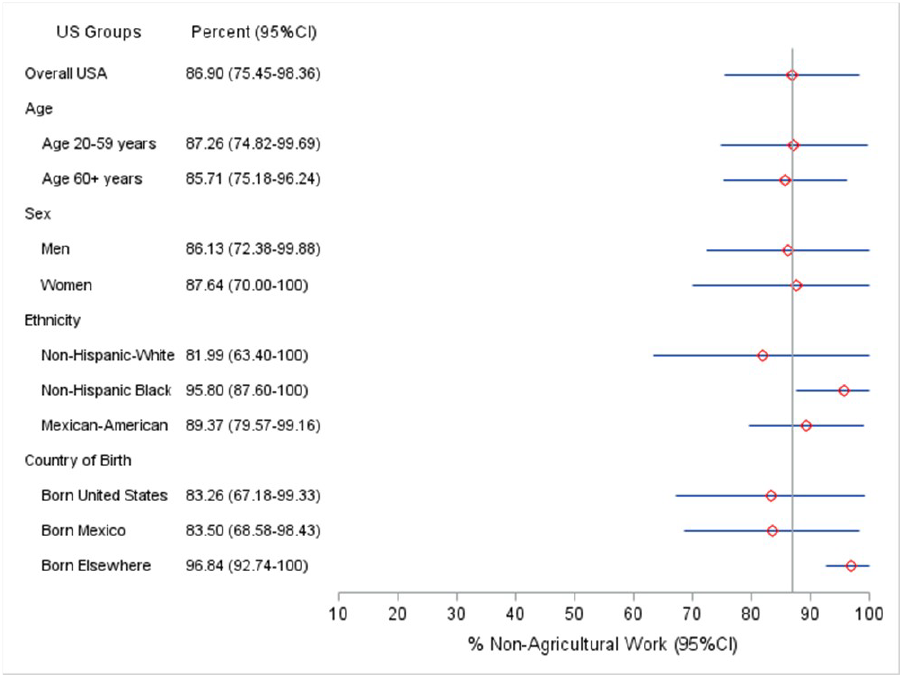
Percent of Seropositives with No History of Agricultural Work.

### 3.4 Length of residence data for US immigrants

Higher rates of positive *C. burnetii* serology in US foreign born residents could potentially reflect infections acquired prior to US immigration. Mexican Americans were a large demographic sub-group and constituted the majority of US immigrants in 2003-04. Because of low subsample sample sizes, most length of residence data for foreign-born Mexican Americans could not be statistically analyzed. However, for perspective, of the 854 Mexican American participants in the sample, an estimated 41% were native born US citizens and 59% were born in Mexico. An estimated 25% of those born in Mexico were naturalized US Citizens; virtually all (96%) of these had resided in the US for 10 or more years and 63% for ≥20 years. Among all Mexican born non-US citizens, an estimated 46% had lived in the US 10 years or more and 19% for ≥20 years. Among seropositive Mexico born non-citizens, an estimated 59% had US residence ≥10 years and 9% ≥ 20 years.

### 3.5 Occupational data analysis

Direct analysis of seropositive risks in the 41 NHANES occupational group job titles was not feasible due to sample size limitations. This problem persisted when the data was further condensed into 17 occupational groups. Supplemental Table 1 shows the crude data distribution for the 17 job categories ordered by the percent *C. burnetii* seropositive. Table 4 shows that when more general level socioeconomic groups were compared, manufacturing, repair, construction, transportation and freight/materials workers had the highest seroprevalence, 3.9% (2.4-5.8) and professional, technical and office workers had the lowest, 2.0% (1.1-3.3). This was a statistically significant difference (t=3.17, p<.01).

**Table 4.**
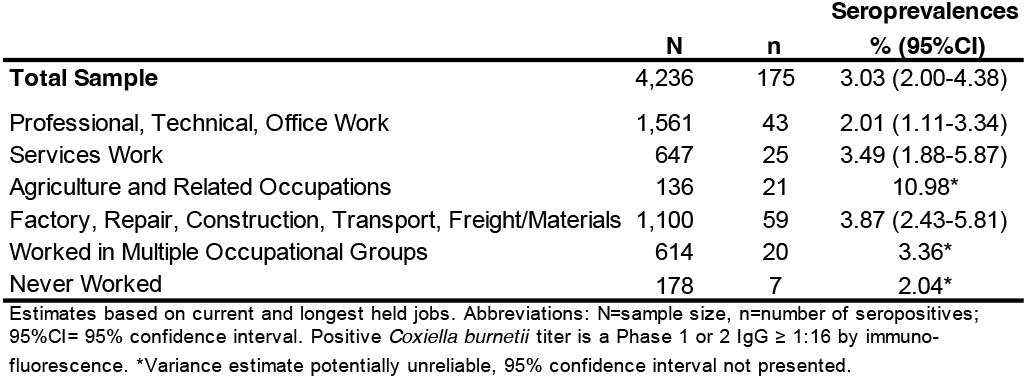
*Coxiella burnetii* Seroprevalences in Major US Occupational Groups.

## 4. Discussion

As initially recognized in the 1950’s, *C. burnetii* infections are endemic in almost all countries, causing disease in both humans and animals [1,26-31). The United States is no exception. A US clinical laboratories serology study, US Q fever notifiable disease reports and the nationally representative US National Inpatient Sample survey document *C. burnetii* infections, Q fever cases and hospitalizations occurring throughout the US [1,32-35]. The NHANES survey seroprevalence data adds further important population-based perspective on US Q fever related exposures.

The NHANES *C. burnetii* seroprevalence results are older data, however the dataset is unique with its large-scale, nationally representative, population-based, in-person active surveillance sampling. Additionally, NHANES 2003-04 had a high survey response rate. The overall and age-specific NHANES 2003-04 estimates for current labor force participation rates in the 20+ years study age range were consistent with official US Bureau of Labor Statistics values, an added indication that the NHANES 2003-04 occupation data are nationally representative (e.g. sampling frame external validation). The 2-year NHANES work history sample size (the total person-years worked) was more than sufficient to estimate prevalences and the percentages of seropositive adults with no history of work in the agricultural sector.

Rather than being rare, *C. burnetii* infection was shown to be common in the US: 3% of the adult population, or some 6.2 million persons, had positive serology. Overall, 87% of those *C. burnetii* seropositive reported no prior history of work in the agricultural sector, equivalent some 5.5 million persons in the US. This finding was consistent across all the major US demographic sub-groups, including US citizens and immigrants. Also, the general-level occupational group analysis here shows that manufacturing, repair, construction, transportation and freight/materials workers in the community may have significant *C. burnetii* exposures. This is consistent with the prior disease outbreaks and case reports.

Collectively the above findings are remarkable as farm worker exposure to livestock is usually considered to be the cause of most US *C. burnetii* infections. Nevertheless, results here suggest that general population exposures to *C. burnetii* may be common and may exceed those of livestock workers. Additional focused studies are needed to more rigorously define the population-level burden of these community level exposures. Table 5 summarizes known and probable *C. burnetii* reservoirs, transmission settings and pathways relevant to community and non-farm related occupational exposures [36-125]. The table emphasizes North American studies so is not globally comprehensive.

**Table 5.**
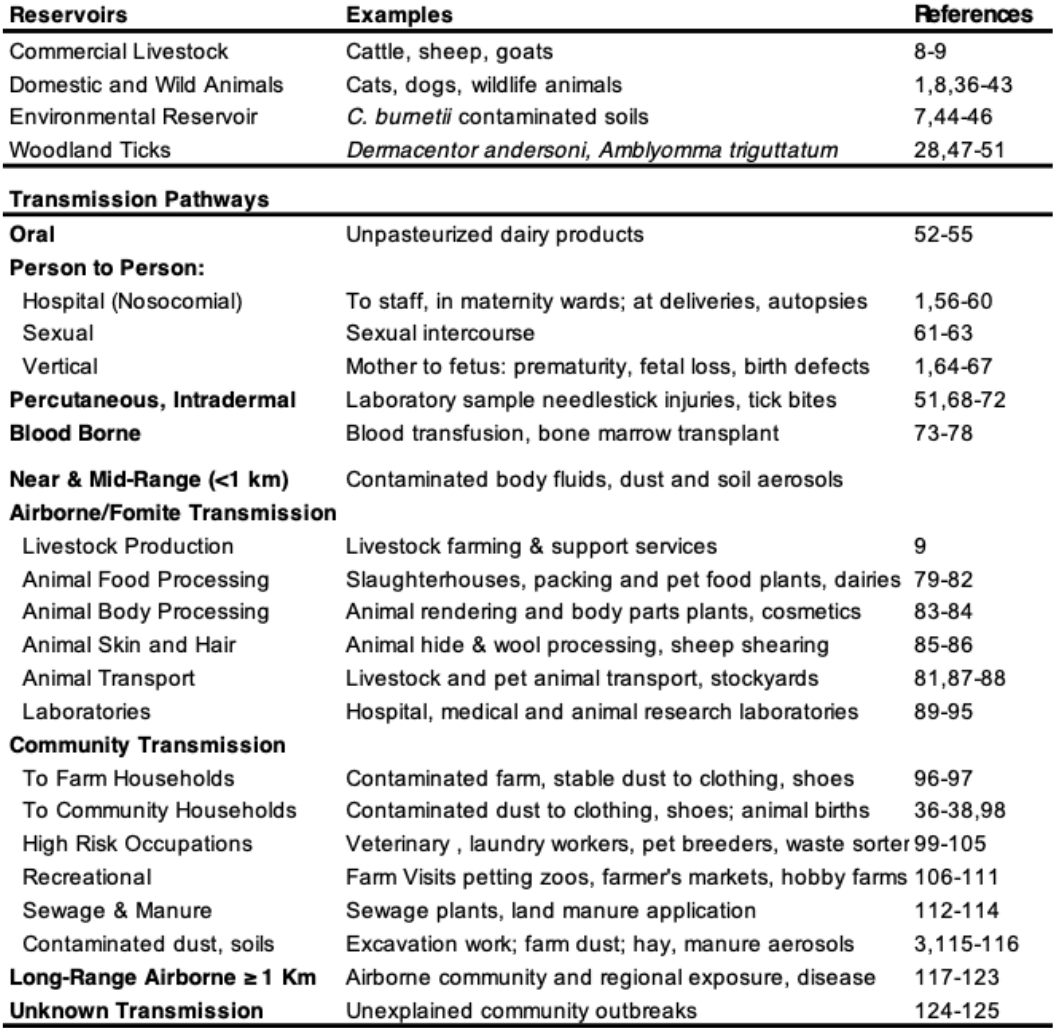
*coxiella burnetii* Reservoirs, Transmission pathways.

| Reservoirs | Examples | References |
| --- | --- | --- |
| Commercial Livestock | Cattle, sheep, goats | 8-9 |
| Domestic and Wild Animals | Cats, dogs, wildlife animals | 1,8,36-43 |
| Environmental Reservoir | *C. burnetii* contaminated soils | 7,44-46 |
| Woodland Ticks | *Dermacentor andersoni*, *Amblyomma triguttatum* | 28,47-51 |
| **Transmission Pathways** |  |  |
| **Oral** | Unpasteurized dairy products | 52-55 |
| **Person to Person:** |  |  |
| Hospital (Nosocomial) | To staff, in maternity wards; at deliveries, autopsies | 1,56-60 |
| Sexual | Sexual intercourse | 61-63 |
| Vertical | Mother to fetus: prematurity, fetal loss, birth defects | 1,64-67 |
| **Percutaneous, Intradermal** | Laboratory sample needlestick injuries, tick bites | 51,68-72 |
| **Blood Borne** | Blood transfusion, bone marrow transplant | 73-78 |
| **Near & Mid-Range (<1 km)** | Contaminated body fluids, dust and soil aerosols |  |
| **Airborne/Fomite Transmission** |  |  |
| Livestock Production | Livestock farming & support services | 9 |
| Animal Food Processing | Slaughterhouses, packing and pet food plants, dairies | 79-82 |
| Animal Body Processing | Animal rendering and body parts plants, cosmetics | 83-84 |
| Animal Skin and Hair | Animal hide & wool processing, sheep shearing | 85-86 |
| Animal Transport | Livestock and pet animal transport, stockyards | 81,87-88 |
| Laboratories | Hospital, medical and animal research laboratories | 89-95 |
| **Community Transmission** |  |  |
| To Farm Households | Contaminated farm, stable dust to clothing, shoes | 96-97 |
| To Community Households | Contaminated dust to clothing, shoes; animal births | 36-38,98 |
| High Risk Occupations | Veterinary, laundry workers, pet breeders, waste sorter | 99-105 |
| Recreational | Farm visits petting zoos, farmer's markets, hobby farms | 106-111 |
| Sewage & Manure | Sewage plants, land manure application | 112-114 |
| Contaminated dust, soils | Excavation work; farm dust; hay, manure aerosols | 3,115-116 |
| **Long-Range Airborne ≥ 1 Km** | Airborne community and regional exposure, disease | 117-123 |
| **Unknown Transmission** | Unexplained community outbreaks | 124-125 |

The results here have some precedents in the literature. In a recent update to the US national NNDSS Q fever case notifications for 2008 to 2017, 60% of officially reported cases had no exposure to animals prior to the onset of their illness and only 40% were employed in high-risk occupations [10]. Also, in the recent large-scale regional Q fever epidemic in the Netherlands, only 3.2% of officially notified Q fever cases had worked in the agriculture sector and 0.5% worked in the meat-processing industry [126-127]. A recent comprehensive global review of Q fever outbreaks showed that half occurred in communities and not in traditional at-risk occupational settings [26]. Of community outbreaks, only half were associated with living in proximity to livestock holdings. Indirect transmission via environmental contamination and airborne spread were the most common infection routes, particularly for large scale urban outbreaks.

Also significantly, a US national-scale environmental survey has demonstrated widespread *C. burnetii* contamination in US dust and soils [7]. Positive samples were found in livestock operations as expected but also in non-agricultural locations such as post offices, retail stores, schools, a bank, a government building and a community center. Given *C. burnetii’s* environmental viability and low infectious dose, aerosolization of pathogenic bacteria from such contaminated dust and soils could pose a significant health hazard to the general public.

### 4.1 Limitations

This study provides a US *C. burnetii* exposure assessment but should not be interpreted as an assessment of the US prevalence of actual clinical disease, e.g. of acute or chronic Q fever. Seroprevalence exposure assessments are typically employed to model disease risks, to identify high risk settings and to characterize vulnerable population subgroups. An example, is to provide guidance for planning preventive vaccination programs. Currently preventive animal and human Q fever vaccines are only available in Australia [31,128-129]. The human vaccine is contraindicated in those with prior *C. burnetii* infection, so pre-vaccination skin testing and serology screening is required. Vaccines are under development to address this limitation as well as to improve immunologic targeting [129].

We could not determine whether foreign born participants had *C. burnetii* exposure in the US or their home country. However, as seropositivity generally wanes with time, especially over decades, the length of immigrant residence in the US provides some initial perspective on whether infection may have occurred pre-immigration or in the US. In the study time frame Mexican Americans were the majority of US immigrants. However a substantial proportion of foreign-born Mexican American immigrants had lived in the US for one or more decades, so a large fraction of these may have been exposed to C. *burnetii* in the US. Also, like native born US citizens, most Mexican American immigrants reported no prior history of work in the agriculture sector, 83.3% vs. 83.5% respectively.

This cross-sectional study employed the NHANES occupational history questionnaire, a robust general-purpose public health surveillance epidemiology instrument. However, it lacks additional detail typically seen in Q fever outbreak investigations. A future specifically designed NHANES study could be fielded to specifically address this limitation. In this regard, there are two biases in the current study that function in opposite directions. First, while the agricultural sector work variable used here is based on the participant’s current as well as longest held job data and captures a significant proportion of an individual’s work history, it does not provide a complete employment history so some prior work in agriculture may not be accounted for. On the other hand, the NHANES survey agricultural work variable available used here is a general-level one that includes both the crop production and the livestock sectors. As levels of US employment solely in the crop production sector are substantial, a significant fraction of those reporting a history of agricultural work in this study had no work contact with livestock [130].

A primary limitation of this study is the retrospective analysis of stored sera data, however this does not reflect on NHANES’s capabilities. Stored sera studies are highly useful as seen here. Nevertheless, NHANES’s primary purpose is to field in-person, designed for purpose, active surveillance public health studies, such as its US national surveillance programs for infectious diseases and environmental exposures monitoring. In any given year, NHANES samples 15 selected US Census tracts. The 2-year sample used here is a minimum required for NHANES analyses. Given an overall 3% *C. burnetii* US seroprevalence, there was reduced study power and ability to provide key subgroup estimates such as infection risks for those working in most occupations including agriculture. A 6-year sample of NHANES stored sera (90 US Census tracts) would be required to address this limitation, and for example, to assess detailed exposure risks in the 17 occupational groups listed in Supplemental Table 1 [131].

## Data Availability

This research was based on publicly available NHANES data. https://www.n.cdc.gov/nchs/nhanes/continuousnhanes/default.aspx?BeginYear=2003

## Supplementary Materials

The following supporting information can be downloaded at: www.mdpi.com/xxx/s1, Table 1: *Coxiella burnetii* Seropositivity by History of Work in 17 Occupational Groups; Table 2: US Bureau of Labor Statistics 2003 Labor Force Participation Rates; Table 3: US Bureau of Labor Statistics 2004 Labor Force Participation Rates; Table 4: Person-Years Worked -Adults Not Currently Employed.

## Author Contributions

Conceptualization, C.D. and M.D.; methodology, C.D and M.D.; software, C.D and G.D.; formal analysis, C.D., G.D and M.D; writing original draft, C.D., G.D and M.D.; writing – review and editing. C.D., G.D and M.D.; visualization, C.D.; project supervision, C.D. All authors have read and agreed to the published version of the manuscript.

## Institutional Review Board Statement

The study was conducted in accordance with the Declaration of Helsinki and approved by the US National Center for Health Statistics Ethics Review Board, Protocol Code #98-12. https://www.cdc.gov/nchs/nhanes/about/erb.html

## Informed Consent Statement

NHANES obtained written informed consent from all subjects involved in the study. This included permission to publish deidentified results in scientific journals and reports. https://www.n.cdc.gov/Nchs/Nhanes/ContinuousNhanes/Documents.aspx?BeginYear=2003

## Acknowledgments

We would like to acknowledge the helpful assistance of Nicholas Schaffer, Economist at the U.S. Bureau of Labor Statistics Division of Information Services for providing the BLS 2003 and 2004 age and sex-specific current US labor force participation data used in this paper. We also thank Dr. Michael Dillon, Senior Scientist at the US Lawrence Livermore National Laboratory for his work in paper conceptual development, data analysis strategy and manuscript development.

## Abbreviations

The following abbreviations are used in this manuscript:

NHANES: US National Health & Nutrition Examination Survey
NCHS: US National Center for Health Statistics
BLS: US Bureau of Labor Statistics
NNDSS: US National Notifiable Diseases Surveillance System
ELISA: Enzyme-linked immunosorbent assay

## Disclaimer/Publisher’s Note

The statements, opinions and data contained in all publications are solely those of the individual author(s) and contributor(s) and not of MDPI and/or the editor(s). MDPI and/or the editor(s) disclaim responsibility for any injury to people or property resulting from any ideas, methods, instructions or products referred to in the content.

## References

Anderson A., Bijlmer H., Fournier P-E., Graves S., Hartzell J., Kersh G.J., Limonard G., Marrie T.J., Massung R.F., McQuiston J. H., et al. Diagnosis and management of Q fever--United States, 2013: recommendations from CDC and the Q fever working group. MMWR Recomm Rep RR-03. 2013, 62, 1–30.

CDC Wonder. Notifiable infectious diseases and conditions, United States: annual tables, 2022. Available online: https://wonder.cdc.gov/nndss/static/2022/annual/2022-table4.html (accessed 30 January 2025).

Parker, N.R., Barralet, J.H., Bell, A.M. Q fever. Lancet. 2006, 367, 679–688. doi: 10.1016/S0140-6736(06)68266-4.

Brooke, R.J., Kretzschmar, M.E., Mutters, N.T., Teunis, S.P. Human dose response relation for airborne exposure to Coxiella burnetii. BMC Infectious Diseases. 2013, 13, 488. doi: 10.1186/1471-2334-13-488.

Heppell, C.W., Egan, J.R., Hall, I. A human time dose response model for Q fever. Epidemics. 2017, 21,30-38. doi: 10.1016/j.epidem.2017.06.001.

Eldin, C., Mélenotte, C., Mediannikov, O., Ghigo, E., Million, M., Edouard, S., Mege, J.L., Maurin, M., Raoult, D. From Q fever to Coxiella burnetii infection: a paradigm change. Clin Microbiol Rev. 2017, 30, 115–190. doi: 10.1128/CMR.00045-16.

Kersh, G.J., Wolfe, T.M., Fitzpatrick, K.A., Candee, A.J., Oliver, L.D., Patterson, N.E., Self, J.S., Priestley, R.A., Loftis, A.D., Massung. R.F. Presence of Coxiella burnetii DNA in the environment of the United States, 2006 to 2008. Appl Environ Microbiol. 2010, 76, 4469–75. doi: 10.1128/AEM.00042-10.

McQuiston, J.H., Childs, J.E. Q fever in humans and animals in the United States. Vector Borne Zoonotic Dis. 2002, 2, 179–91 (2002). doi: 10.1089/15303660260613747.

National Association of State Public Health Veterinarians, National Assembly of State Animal Health Officials. Prevention and control of Coxiella burnetii infection among humans and animals: guidance for a coordinated public health and animal health response. Available online: http://nasphv.org/Documents/Q_Fever_2013.pdf (accessed 2 Febuary 2025).

Cherry, C.C., Heitman K.N., Bestul, N.C., Kersh, G.J. Acute and chronic Q fever national surveillance - United States, 2008-2017. Zoonoses Public Health. 2022, 69, 73–82 (2022). doi: 10.1111/zph.12896.

Anderson, A.D., Kruszon-Moran, D., Loftis, A.D., McQuillan, G., Nicholson, W.L., Priestley, R.A., Candee, A.J., Patterson, N.E., Massung R.F. Seroprevalence of Q fever in the United States, 2003-2004. Am J Trop Med Hyg. 2009, 81, 691–4. doi: 10.4269/ajtmh.2009.09-0168.

US National Health and Nutrition Examination Survey. Questionnaire: SP (2003-04) occupation. OCQ-C. Available online: https://www.n.cdc.gov/nchs/data/nhanes/public/2003/questionnaires/sp_ocq_c.pdf (accessed 2 February 2025).

Zipf, G., Chiappa, M., Porter, K.S., Ostchega, Y., Lewis, B.G., Dostal, J. National Health and Nutrition Examination Survey: plan and operations, 1999-2010. Vital Health Stat 1. 2013, 56,1-28.

Curtin, L.R., Mohadjer, L.K., Dohrmann, S.M., Montaquila J.M., Kruszan-Moran, D., Mirel, L.B., Carroll, M.D., Hirsch, R., Schober, S., Johnson, C.L. The National Health and Nutrition Examination Survey: sample design, 1999-2006. Vital Health Stat 2. 2012, 155, 1–39.

US National Health and Nutrition Examination Survey. Unweighted response rates for NHANES 2003-2004 by age and gender. Available online: https://www.n.cdc.gov/nchs/data/ResponseRates/RRT0102MF.pdf (accessed 2 February 2025).

US National Health and Nutrition Examination Survey. 2003-2004 Data documentation, codebook, and frequencies occupation (OCQ_C). Available online: https://www.n.cdc.gov/Nchs/Nhanes/2003-2004/OCQ_C.htm (accessed 2 February 2025).

National Institute for Occupational Safety and Health. Collecting and using industry and occupation data. Available on-line: https://www.cdc.gov/niosh/occupation-industry-data/about-data/coding/classification-systems.html?CDC_AAref_Val= **https://www.cdc.gov/niosh/topics/coding/more.htmlhtml (accessed 2 February 2025).

Krieger, N., Bareau, E.M., Soobader, M.J. Class matters: U.S. versus U.K. measures of occupational disparities in access to health services and health status in the 2000 U.S. National Health Interview Survey. Int J Health Serv. 2005, 35, 213–36. doi: 10.2190/JKRE-AH92-EDV8-VHYC.

U. S. Bureau of Labor Statistics. Labor Force Statistics from the Current Population Survey. Concepts and Definitions. Available online: https://www.bls.gov/cps/definitions.htm (accessed 2 February 2025).

US National Health and Nutrition Examination Survey. 2003-2004 data documentation, codebook, and frequencies. Coxiella burnetii (Q fever) antibodies - serum (surplus) (SSQFEV_C), update 2022. Available online: https://www.n.cdc.gov/nchs/nhanes/search/datapage.aspx?Component=Laboratory&CycleBeginYear=2003 (accessed 2 February 2025).

Péter, O., Dupuis, G., Burgdorfer, W., Peacock, M. Evaluation of the complement fixation and indirect immunofluorescence tests in the early diagnosis of primary Q fever. Eur J Clin Microbiol. 1985, 4, 394–396. doi: 10.1007/BF02148690

Philip, R.N., Casper, E.A., Ormsbee, R.A., Burgdorfer, W. Microimmunofluorescence test for the serological study of rocky mountain spotted fever and typhus. J Clin Microbiol. 1976, 3, 51–61 (1976). doi: 10.1128/jcm.3.1.51-61.1976.

Parker, J.D., Talih, M., Irimata, K.E., Zhang, G., Branum, A.M., Davis, D., Das, B., Hamilton, B.E., Kochanek, K. National Center for Health Statistics data presentation standards for rates and counts. Vital Health Stat 2. 2023, 200. Available online: https://stacks.cdc.gov/view/cdc/124368. (accessed 30 January 2025).

Parker J.D., Talih, M., Malec, D.J., Beresovsky. V., Carroll, M., Gonzalez, J.F., Hamilton, B.E., Ingram, D.D., Kochanek, K., McCarty, F. National Center for Health Statistics data presentation standards for proportions. Vital Health Stat 2. 2017, 175, 1–22. Available online: https://www.cdc.gov/nchs/data/series/sr_02/sr02_175.pdf (accessed 2 February 2025).

US National Health and Nutrition Examination Survey. NHANES tutorials, reliability of estimates. Available online: https://www.n.cdc.gov/nchs/nhanes/tutorials/ReliabilityOfEstimates.aspx (accessed 2 February 2025).

Tan, T., Heller, J., Firestone, S., Stevenston, M., Wiethoelter, A. A systematic review of global Q fever outbreaks. One Health. 2023, 18, 100667. doi: 10.1016/j.onehlt.2023.100667.

Ahaduzzaman, M., Reza, M.M. Global and regional seroprevalence of coxiellosis in small ruminants: a systematic review and meta-analysis. Veterinary Medicine and Science. 2024, 10, e1441. doi: 10.1002/vms3.1441.

Berge, T.O., Lennette, E.H. World distribution of Q fever: human, animal and arthropod infection. Am J Hyg. 1953, 57, 125–143.

Gisbert, P., Garcia-Ispierto, I., Quintela, L.A., Guatteo, R. Coxiella burnetii and reproductive disorders in cattle: A Systematic Review. Animals (Basel). 2024, 14, 1313. doi: 10.3390/ani14091313.

Olivas, S. et al. Massive dispersal of Coxiella burnetii among cattle across the United States. Microb Genom. 2016, 2, e000068. doi: 10.1099/mgen.0.000068.

Toledo-Perona, R., Contreras, A., Gomis, J., Quereda, J.J., García-Galán, A., Sánchez, A, Gómez-Martín, Á. Controlling Coxiella burnetii in naturally infected sheep, goats and cows, and public health implications: a scoping review. Front Vet Sci. 2024, 11, 1321553. doi: 10.3390/ani14091313.

Agency for Healthcare Quality and Research. Overview of the National (nationwide) Inpatient Sample (NIS). Available online: https://hcup-us.ahrq.gov/nisoverview.jsp (accessed 2 February 2025).

Centers for Disease Control and Prevention. Q fever epidemiology and statistics. Available online: https://www.cdc.-gov/q-fever/data-research/index.html (accessed 2 Febuary 2024).

Miller HK, Binder AM, Peterson A, Theel ES, Volpe JM, Couturier MR, Cherry CC, Kersh GJ. Trends in Q fever serologic testing by immunofluorescence from four large reference laboratories in the United States, 2012-2016. Sci Rep. 2018, 8, 16670. doi: 10.1038/s41598-018-34702-2.

Mohamad Alahmad, M.A., Hammoud, K.A. Inpatient Q fever frequency is on the rise. Can J Infect Dis Med Microbiol. 2023, 31, 4243312. doi: 10.1155/2023/4243312.

Ebani, V.V. Coxiella burnetii Infection in Cats. Pathogens. 2023, 12, 1415. doi: 10.3390/pathogens12121415

Pinsky, R.L., Fishbein, D.B., Greene, C.R., Gensheimer, K.F. An outbreak of cat-associated Q fever in the United States. J Infect Dis. 1991, 164, 202–4. doi: 10.1093/infdis/164.1.202

Langley JM, Marrie TJ, Covert A, Waag DM, Williams JC. Poker Players’ Pneumonia: An urban outbreak of Q fever following exposure to a parturient cat. N Engl J Med. 1988, 319, 6–. doi: 10.1056/NEJM198808113190607

Marrie, T.J., Langille, D., Papukna, V., Yates, L. Truckin’ pneumonia–an outbreak of Q fever in a truck repair plant probably due to aerosols from clothing contaminated by contact with newborn kittens. Epidemiol Infect. 1989, 102, 127–. doi: 10.1017/s0950268800029757

Buhariwalla, F., Cann, B., Marriem T.J. A dog-related outbreak of Q fever. Clin Infect Dis. 1996, 23, 753–5. doi: 10.1093/clinids/23.4.753

McMillan, I.A., Norris, M.H., Golon, S.J., Franckowiak, G.A., Grinolds, J.M., Goldstein, S.M., Phelps, D.M., Bodenchuk, M.J., Leland, B.R., Bowen, R.A., et al. Serosurveillance of Coxiella burnetii in feral swine populations of Hawai’i and Texas identifies overlap with human Q fever incidence. J Clin Microbiol. 2024. Aug 27:e0078024. doi: 10.1128/jcm.00780-24.

Reusken C, van der Plaats R, Opsteegh M, de Bruin, A., Swart, A. Coxiella burnetii (Q fever) in Rattus norvegicus and Rattus rattus at livestock farms and urban locations in the Netherlands; could Rattus spp. represent reservoirs for (re)introduction? Prev Vet Med. 2011, 101:124–30. doi: 10.1016/j.prevetmed.2011.05.003

Meredith, A., Cleaveland, S.C., Denwood, M.J., Brown, J.K., Shaw, D.J. Coxiella burnetii (Q-Fever) Seroprevalence in Prey and Predators in the United Kingdom: Evaluation of Infection in Wild Rodents, Foxes and Domestic Cats Using a Modified ELISA. Transbound Emerg Dis. 2015. 62, 639–49. doi: 10.1111/tbed.12211

Kersh GJ, Fitzpatrick KA, Self JS, Priestley RA, Kelly AJ, Lash RR, Marsden-Haug N, Nett RJ, Bjork A, Massung RF, et al. Presence and persistence of Coxiella burnetii in the environments of goat farms associated with a Q fever outbreak. Applied and Environmental Microbiology. 2013, 79, 1703–. 10.1128/AEM.03472-12

de Bruin, A., van der Plaats, R.Q., de Heer, L., Paauwe, R., Schimmer, B., Vellema, P., van Rotterdam, B.J., van Duynhoven, Y.T. Detection of Coxiella burnetii DNA on Small-Ruminant Farms during a Q fever Outbreak in the Netherlands. Applied and Environmental Microbiology. 2012, 78, 1657–. doi: 10.1128/AEM.07323-11

de Bruin, A., Janse, I., Koning, M., de Heer, L., van der Plaats, R.Q,. van Leuken, J.P., van Rotterdam, B.J. Detection of Coxiella burnetii DNA in the environment during and after a large Q fever epidemic in the Netherlands. J Appl Microbiol. 2013, 114, 1404–. 10.1111/jam.12163

Yessinou, R.E., Katja, M.S., Heinrich, N., Farougou S. Prevalence of Coxiella-infections in ticks - review and meta-analysis. Ticks Tick Borne Dis. 2022, 13, 101926. doi: 10.1016/j.ttbdis.2022.101926

Duron, O., Sidi-Boumedine, K., Rousset, E., Moutaillerm S., Jourdain, E. The Importance of Ticks in Q fever Transmission: What Has (and Has Not) Been Demonstrated? Trends Parasitol. 2015, 31. 536–552. doi: 10.1016/j.pt.2015.06.014

Graves, S.R., Gerrard, J., Coghill, S. Q fever following a tick bite. Aust J Gen Pract. 2020. 49, 823–825. doi: 10.31128/AJGP-01-20-5195

Beaman, M.H., Hung, J. Pericarditis associated with tick-borne Q fever. Aust N Z J Med. 1989, 19, 254–6. doi: 10.1111/j.1445-5994.1989.tb00258

Nett RJ, Book E, Anderson AD. Q fever with unusual exposure history: a classic presentation of a commonly misdiagnosed disease. Case Rep Infect Dis. 2012, 916142. doi: 10.1155/2012/916142

Kim, S.G., Kim, E.H., Lafferty, C.J., Dubovi, E. Coxiella burnetii in bulk tank milk samples, United States. Emerg Infect Dis. 2005, 11, 619–21. doi: 10.3201/eid1104.041036

Signs, K.A., Stobierski, M.G., Gandhi, T.N. Q fever cluster among raw milk drinkers in Michigan, 2011. Clin Infect Dis. 2012, 55, 1387–9. doi: 10.1093/cid/cis690

Beck, M.D., Bell, J.A., Shaw, E.W., Heubner, R.J. Q fever studies in Southern California; an epidemiological study of 300 cases. Public Health Rep. 1949, 14, 41–56.

Gale, P., Kelly, L., Mearns, R., Duggan, J., Snary E.L. Q fever through consumption of unpasteurised milk and milk products - a risk profile and exposure assessment. J Appl Microbiol. 2015, 118, 1083–95. doi: 10.1111/jam.12778

Deutsch, D.L., Peterson, E.T. Q fever: Transmission from one human being to others. J Am Med Assoc. 1950, 143, 348–50. doi: 10.1001/jama.1950.02910390020006

Amit, S., Shinar, S., Halutz, O., Atiya-Nasagi, Y., Giladi, M. Suspected person-to-person transmission of Q fever among hospitalized pregnant women. Clin Infect Dis. 2014, 58:e, 146–7. doi: 10.1093/cid/ciu151

Raoult, D., Stein, A. Q fever during pregnancy--a risk for women, fetuses, and obstetricians. N Engl J Med. 1994, 330, 371. doi: 10.1056/nejm199402033300518

Harman, J.B. Q fever in Great Britain; clinical account of eight cases. Lancet. 1949, 2(6588), 1028–30. doi: 10.1016/s0140-6736(49)91600-1

Osorio, S., Sarriá, C., González-Ruano, P/, Casal, E.C., García, A. Nosocomial transmission of Q fever. Journal of Hospital Infection. 2003, 54, 162–163. 10.1016/S0195-6701(03)00111-7

Milazzo, A., Hall, R., Storm, P.A., Harris, R.J., Winslow, W., Marmion, B.P. Sexually transmitted Q fever. Clin Infect Dis. 2001, 33, 399–402. doi: 10.1086/321878

Kruszewska, D., Lembowicz, K., Tylewska-Wierzbanowska, S. Possible sexual transmission of Q fever among humans. Clin Infect Dis. 1996, 22, 1087–8. doi: 10.1093/clinids/22.6.1087

Miceli, M.H., Veryser, A.K., Anderson, A.D., Hofinger, D., Lee, S.A., Tancik, C. A case of person-to-person transmission of Q fever from an active duty serviceman to his spouse. Vector-Borne and Zoonotic Diseases. 2010, 10, 539–41. doi: 10.1089/vbz.2009.0101

Raoult D, Stein A. Q fever during pregnancy--a risk for women, fetuses, and obstetricians. N Engl J Med. 1994, 330, 371. doi: 10.1056/nejm199402033300518

Million, M., Roblot, F., Carles, D., D’Amato, F., Protopopescu, C., Carrieri, M.P., Raoult, D. Reevaluation of the risk of fetal death and malformation after Q fever. Clin Infect Dis. 2014, 59, 256–60. doi: 10.1093/cid/ciu259

Levin, G., Herzberg, S., Attari, R., Abu Khatab, A., Gil. M., Rottenstreich, A. Q fever first presenting as a septic shock resulting in intrauterine fetal death. Eur J Obstet Gynecol Reprod Biol. 2018, 229, 204–205. doi: 10.1016/j.ejogrb.2018.08.021.

Ellis, M.E., Smith, C.C., Moffat, M.A. Chronic or fatal Q-fever infection: a review of 16 patients seen in North-East Scotland (1967-80). Q J Med. 1983, 52, 54–66.

Johnson, J.E., Kadull, P.J. Laboratory-acquired Q fever. A report of fifty cases. Am J Med. 1966, 41, 391–403. doi: 10.1016/0002-9343(66)90085-4

Fonseca, F., Pinto, M.R., De Azevedo, J,F,, Lacerda, M.T. Q Fever in Portugal. History, Clinical Observations, Diagnosis. Clinica Contemporanea. 1949, 3(21, 22, 23), 1159–71. Available online: https://www.cabdirect.org/cabdirect/abstract/19502902290 (accessed 2 February 2025).

Commentary: Experimental Q Fever in Man. British Medical Journal 1950, p. 1000.

Robyn, M.P., Newman, A.P., Amato, M., Walawander, M., Kothe, C., Nerone, J.D., Pomerantz, C., Behravesh, C.B., Biggs, HM., Dahlgren, F.S. et al. Q fever outbreak among travelers to Germany associated with live cell therapy - United States and Canada, 2014: a co-publication. Can Commun Dis Rep. 2015, 41, 223–226. doi: 10.14745/ccdr.v41i10a01

van Kraaij, M.G., Slot, E., Hogema, B.M., Zaaijer, H.L. Lookback procedures after postdonation notifications during a Q fever outbreak in the Netherlands. Transfusion. 2013, 53, 716–21. doi: 10.1111/j.1537-2995.2012.03792.x

Kersh, G.J., Priestley, R., Massung, R.F. Stability of Coxiella burnetii in stored human blood. Transfusion. 2013. 53 1493–6. doi: 10.1111/j.1537-2995.2012.03912.x

Goldberg, J.S., Perkins, H.A., Zapitz, V.M., Hurst, B.B., Suther, D., Jensen, F., Lenette, E.H., Damus, K., Roberto, R.R. Q fever–California. MMWR. Morbidity and Mortality Weekly Report, 1977, 26, 86–91.

B.M., Slot, E., Molier, M., Schneeberger, P.M., Hermans, M.H, van Hannen, E.J., van der Hoek, W., Cuijpers, H.T., Zaaijer, H.L. Coxiella burnetii infection among blood donors during the 2009 Q-fever outbreak in The Netherlands. Transfusion. 2012, 52, 144–50. doi: 10.1111/j.1537-2995.2011.03250.x

Oei, W., Kretzschmar, M.E., Zaaijer, H.L., Coutinho, R., van der Poel, C.L., Janssen, M.P. Estimating the transfusion transmission risk of Q fever. Transfusion. 2014, 54, 1705–11. doi: 10.1111/trf.12539

Kanfer, E., Farrag, N., Price, C., MacDonald, D., Coleman, J., Barrett, A.J. Q fever following bone marrow transplantation. Bone Marrow Transplant. 1988, 3, 165–6

Gürtler, L., Bauerfeind, U., Blümel, J., Burger, R., Drosten, C., Gröner, A., Heiden, M., Hildebrandt, M., Jansen, B., Offergeld, R. et al. Coxiella burnetii - Pathogenic Agent of Q (Query) Fever. 2014. Transfus Med Hemother. 2014, 41, 60–72. doi: 10.1159/000357107

Derrick, E.H. Q Fever a new fever entity: clinical features. diagnosis, and laboratory investigation. Medical Journal of Australia. 1937, 2, 299–. doi:10.5694/j.1326-5377.1937.tb43743.x

Bell, J.A., Beck, M.D., Huebner, R.J. Epidemiologic Studies of Q fever in Southern California. JAMA. 1950, 142, 872–. doi:10.1001/jama.1950.02910300006002

Topping, N.H., Shepard, C.C., Irons, J.V. Q fever in the United States: Epidemiologic Studies of an Outbreak Among Stock Handlers and Slaughterhouse Workers. JAMA. 1947, 133, 815–. doi:10.1001/jama.1947.02880120001001

Uren AM, Harris J, Slinko V, Vosti F, Young M. Q fever infection is a preventable risk associated with pet food manufac^-^ turing. Ann Work Expo Health. 2024, 68, 104–107. doi: 10.1093/annweh/wxad068

National Institute for Occupational Safety and Health (NIOSH). Occupational Hazard Assessment: Criteria for Controlling Occupational Hazards Animal Rendering Processes. Available online: https://www.cdc.gov/niosh/docs/81-133/default.html (accessed 2 February 2025).

Wade, A.J., Cheng, A.C., Athan, E., Molloy, J., Harris, O.C., Stenos, J., Hughes, A.J. Q fever outbreak at a cosmetics supply factory. Clin Infect Dis. 2006. 42(7):e50–2. doi: 10.1086/501127

Wattiau, P., Boldisova, E., Toman, R., Van Esbroeck, M., Quoilin, S., Hammadi S, Tissot-Dupont, H., Raoult, D., Henkinbrandt, J.M. Q fever in Woolsorters, Belgium. Emerg Infect Dis. 2011. 17, 2368–9. doi: 10.3201/eid1712.101786

Sigel MM, Scott TF, Henle W, Janton, O.H. Q fever in a wool and hair processing plant. Am J Public Health Nations Health. 1950, 40, 524–32. doi: 10.2105/ajph.40.5.524

Clark, W.H., Lennette, E.H., Romer, M.S. Q fever in California. IX. An outbreak aboard a ship transporting goats. Am J Hyg. 1951, 54, 35–43.

Alonso, E., Eizaguirre, D., Lopez-Etxaniz, I., Olaizola, J.I., Ocabo, B., Barandika, J.F., Jado, I., Álvarez-Alonso, R., Hurtado, A., García-Pérez, A. et al. A Q fever outbreak associated to courier transport of pets. PLoS One. 2019. 14, (11):e0225605. doi: 10.1371/journal.pone.0225605

Bayer RA. Q fever as an occupational illness at the National Institutes of Health. Public Health Rep. 1982, 97, 58–60.

Hall, C.J., Richmond, S.J., Caul, E.O., Pearce, N.H., Silver, I.A. Laboratory outbreak of Q fever acquired from sheep. Lancet. 1982, 1, (8279):1004–6. doi: 10.1016/s0140-6736(82)92001-3

Meiklejohn, G., Reimer, L.G., Graves, P.S, Helmick, C. Cryptic Epidemic of Q fever in a Medical School. Infect Dis. 1981, 144, 113–, 10.1093/infdis/144.2.107

Galganski, L.A., Keller, B.A., Long, C., Yamashiro, K.J., Hegazi, M.S., Pivetti, C.D., Talken, L.A., Raff, G.W., Farmer, D.L., Chomel, B.B., Ma, B. Minimizing the risk of occupational Q fever exposure: A protocol for ensuring Coxiella burnetii-negative pregnant ewes are used for medical research. Lab Anim. 2021, 55, 170–176. doi: 10.1177/0023677220965628.

Simor, A.E., Brunton, J.L., Salit I.E., Vellend, H., Ford-Jones, L., Spence, L.P. Q fever: hazard from sheep used in research. Can Med Assoc J. 1984, 130, 1013–6.

Curet, L.B., Paust, J.C. Transmission of Q fever from experimental sheep to laboratory personnel. Am J Obstet Gynecol. 1972, 114, 566–68. doi: 10.1016/0002-9378(72)90222-0

Centers for Disease Control. Q fever at a university research center-California. Morbidity Mortality Weekly Rep. 1979, 28, 333–3.

Mann, J.S., Douglas, J.G., Inglis, J.M., Leitch, A.G. Q fever: person to person transmission within a family. Thorax. 1986, 41, 974–5. doi: 10.1136/thx.41.12.974

Schimmer, B., Lenferink, A., Schneeberger, P., Aangenend, H., Vellema, P., Hautvast, J., van Duynhoven, Y. Seroprevalence and risk factors for Coxiella burnetii (Q fever) seropositivity in dairy goat farmers’ households in The Netherlands, 2009–2010. PLoS One. 2012. 7(7)e42364. doi:10.1371/journal.pone.0042364

Proboste, T., Clark, N.J., Tozer. S., Wood, C., Lambert, S.B., Soares Magalhães, R.J. Profiling risk factors for household and community spatiotemporal clusters of Q fever notifications in Queensland between 2002 and 2017. Pathogens. 2022. 11, 830.6. doi: 10.3390/pathogens11080830

Whitney, E.A., Massung, R.F., Candee, A.J., Ailes, E.C., Myers, L.M., Patterson, N.E., Berkelman, R.L. Seroepidemiologic and occupational risk survey for Coxiella burnetii antibodies among US veterinarians. Clin Infect Dis. 2009. 48, 550–7. doi: 10.1086/5967

Vest, K.G., Clark, L.L. Serosurvey and observational study of US Army Veterinary Corps officers for Q fever antibodies from 1989 to 2008. Zoonoses Public Health. 2014, 61, 271–82. doi: 10.1111/zph.12067

Clark, W.H., Bogucki, A.S., Lenette, E.H. Q fever in California. VI. Description of an epidemic occurring at Davis, California, in 1948. Am J Hyg. 1951, 54, 15–24.

Oliphant JW, Gordon DA, Meis, Armon, Parker, RR. Q fever in laundry workers, presumably transmitted from contaminated clothing. Am J Hyg. 1949, 49, 76–82. doi: 10.1093/oxfordjournals.aje.a119261

Shapiro, A.J., Bosward, K.L., Heller, J., Norris, J.M. Seroprevalence of Coxiella burnetii in domesticated and feral cats in eastern Australia. Vet Microbiol. 2015. 177, 154–61. doi: 10.1016/j.vetmic.2015.02.011

Shapiro, A.J., Norris, J.M., Bosward, K.L., Heller, J. Q fever (Coxiella burnetii) Knowledge and Attitudes of Australian Cat Breeders and Their Husbandry Practices. Zoonoses Public Health. 2017, 64, 252–261. doi: 10.1111/zph.12305

Alonso, E., Lopez-Etxaniz, I., Hurtado, A., Liendo, P., Urbaneja, F., Aspiritxaga, I., Olaizola, J.I., Piñero, A., Arrazola, I., Barandika, J.F. Q fever Outbreak among Workers at a Waste-Sorting Plant. PLoS ONE, 2015, 10, (9): e0138817. 10.1371/journal.pone.0138817

Bjork, A., Marsden-Haug, N., Nett, R.J., Kersh, G.J., Nicholson, W., Gibson, D., Szymanski, T., Emery, M., Kohrs, P., Woodhall, D. First reported multistate human Q fever outbreak in the United States, 2011. Vector Borne Zoonotic Dis. 2014 14, 111–7. doi: 10.1089/vbz.2012.1202

Whelan, J., Schimmer, B., de Bruin, A., van Beest Holle, M.R., van der Hoek, W., ter Schegget, R. Visits on ‘lamb-viewing days’ at a sheep farm open to the public was a risk factor for Q fever in 2009. Epidemiology and Infection. 2012, 140, 864–. 10.1017/S0950268811001427

Valkenburgh, S.M., de Bruin, A., Züchner, L. Q-koorts op kinderboerderijen [Q fever on petting zoos]. Tijdschr Dierge-neeskd. 2011. 136, 158–61.

Porten, K., Rissland, J., Tigges, A., Broll, S., Hopp, W., Lunemann, M., van Treeck, U., Kimmig, P., Brockmann, S.O., Wagner-Wiening, A super-spreading ewe infects hundreds with Q fever at a farmers’ market in Germany. BMC Infect Dis. 2006, 6, 147. doi: 10.1186/1471-2334-6-147

Tan, T.S., Hernandez-Jover, M., Hayes, L.M., Wiethoelter, A.K., Firestone, S.M., Stevenson, M.A., Heller, J. Identifying scenarios and risk factors for Q fever outbreaks using qualitative analysis of expert opinion. Zoonoses Public Health. 2022. 69, 344–358. doi: 10.1111/zph

Hurtado, A., Zendoia, I.I., Alonso, E., Beraza, X., Bidaurrazaga, J., Ocabo, B., Arrazola, I., Cevidanes, A., Barandika, J.F., García-Pérez, A.L. A Q fever outbreak among visitors to a natural cave, Bizkaia, Spain, December 2020 to October 2021. Euro Surveill. 2023, 28, 2200824. doi: 10.2807/1560.

Schets, F.M., de Heer, L., de Roda Husman, A.M. Coxiella burnetii in sewage water at sewage water treatment plants in a Q fever epidemic area. Int J Hyg Environ Health. 2013. 216, 98–702. doi: 10.1016/j.ijheh.2012.12.010

Hermans, T., Jeurissen, L., Hackert, V., Hoebe, C. Land-applied goat manure as a source of human Q-fever in the Netherlands, 2006–2010. PLoS One. 2014, 9, (5):e96607. doi: 10.1371/journal.pone.0096607

van den Brom, R., Roest, H.J., de Bruin, A., Dercksen, D., Santman-Berends, I., van der Hoek, W., Dinkla, A., Vellema, J., Vellema, P. A probably minor role for land-applied goat manure in the transmission of Coxiella burnetii to humans in the 2007-2010 Dutch Q fever outbreak. PLoS One. 2015, 27, 10(3):e0121355. doi: 10.1371/journal.pone.0121355

Price, C., Smith, S., Stewart, J., Palesy, T., Corbitt, M., Galappaththy, C., Hanson, J. Increased recognition of Q fever aortitis as a chronic manifestation of Q fever in tropical North Queensland, Australia. Eur J Clin Microbiol Infect Dis. 2023, 42, 1537–1541. doi: 10.1007/s10096-023-04687-6

Salmon, M.M., Howells, B., Glencross, E.J., Evans, A.D., Palmer, S.R. Q fever in an urban area. Lancet. 1982, 1(8279), 1002–4. doi: 10.1016/s0140-6736(82)92000-1.

Dillon, C.F., Dillon, M.B. Multi-Scale Airborne Infectious Disease Transmission. Appl Environ Microbiol. 2021, 87, e02314–20. doi: 10.1128/AEM.02314-20

Dillon, M.B., Dillon, C.F. Regional Relative Risk, a Physics-Based Metric for Characterizing Airborne Infectious Disease Transmission. Appl Environ Microbiol. 2021, 87, (21):e0126221. doi: 10.1128/AEM.01262-21

Gilsdorf, A., Kroh, C., Grimm, S., Jensen, E., Wagner-Wiening, C., Alpers. K. Large Q fever outbreak due to sheep farming near residential areas, Germany, 2005. Epidemiol Infect. 2008, 136, 1084–7. doi: 10.1017/S0950268807009533

Hawker, J.I., Ayres, J.G., Blair, I., Evans, M.R., Smith, D.L., Smith, E.G., Burge, P.S., Carpenter, M.J., Caul, E.O., Coupland, B., et al. A large outbreak of Q fever in the West Midlands: windborne spread into a metropolitan area? Commun Dis Public Health. 1988, 1, 180–187.

Tissot-Dupont, H., Torres, S., Nezri, M., Raoult, D. Hyperendemic focus of Q fever related to sheep and wind. Am J Epidemiol. 1999, 50, 67–74. doi: 10.1093/oxfordjournals.aje.a009920

Hackert, V.H., van der Hoek, W., Dukers-Muijrers, N., de Bruin, A., Al Dahouk, S., Neubauer, H., Bruggeman, C.A., Hoebe, C.J. et al. Q fever: Single-Point Source Outbreak with High Attack Rates and Massive Numbers of Undetected Infections Across an Entire Region. Clinical Infectious Diseases. 2012, 55, 1591–1599. doi:10.1093/cid/cis734

Hackert, V.H., Hoebe, C.J.P.A., Dukers-Muijrers, N., Krafftm T., Kauh, l B., Henning, K., Karges, W., Sprague, L., Neubauer, H., Al Dahouk, S. et al. Q fever: Evidence of a Massive Yet Undetected Cross-Border Outbreak, with Ongoing Risk of Extra Mortality, in a Dutch-German Border Region. Transbound Emerg Dis. 2020. 67, 1660–1670. doi:10.1111/tbed.13505

Clark, W.H., Romer, M.S., Holmes, M.A., Welsh, H.H., Lennette, E.H., Abinanti, F.R. Q fever in California VIII. An epidemic of Q fever in a small rural community in northern California. Am J Hyg. 1951, 54, 25–34.

Archer, B.N., Hallahan, C., Stanley, P., Seward, K., Lesjak, M., Hope, K., Brown, A. Atypical outbreak of Q fever affecting low-risk residents of a remote rural town in New South Wales. Commun Dis Intell Q Rep. 2017, 41, E125–E133.

van der Hoek, W, Morroy, G., Renders, N.H., Wever, P.C., Hermans, M.H., Leenders, A.C., Schneeberger, P.M. Epidemic Q fever in humans in the Netherlands. Adv Exp Med Biol. 2012, 984, 329–64. doi: 10.1007/978-94-007-4315-1_17

Dijkstra, F., van der Hoek, W., Wijers, N., Schimmer, B., Rietveld, A., Wijkmans, CJ, Vellema, P., Schneeberger, P.M. The 2007–2010 Q fever epidemic in the Netherlands: characteristics of notified acute Q fever patients and the association with dairy goat farming. FEMS Immunol Med Microbiol. 2012, 64, 12–. doi: 10.1111/j.1574-695X.2011.00876.x.

O’Neill, T.J., Sargeant, J.M., Poljak, Z. The effectiveness of Coxiella burnetii vaccines in occupationally exposed populations: a systematic review and meta-analysis. Zoonoses Public Health. 2014, 61, 81–96. doi: 10.1111/zph.12054.

Sam, G., Stenos, J., Graves, S.R., Rehm, B.H.A. Q fever immunology: the quest for a safe and effective vaccine. NPJ Vaccines. 2023, 8, 133. (2023). doi: 10.1038/s41541-023-00727-6.

US Department of Agriculture Economic Research Service. Farm labor (see U.S. employment in agriculture and support industries, 2001-23). Available online: https://www.ers.usda.gov/topics/farm-economy/farm-labor#size (accessed 2 February 2025).

Johnson, C.L., Paulose-Ram, R., Ogden, C.L., Carroll, M.D., Kruszon-Moran, D., Dohrmann, S.M., Curtin, L.R. National Health and Nutrition Examination Survey: Analytic Guidelines, 1999-2010. Vital Health Stat 2. 2013, 161, 1–24.

