## Supplementary Materials for "Q Fever Related Community Infections, US Exposure to *Coxiella burnetii*"

**Community Exposure to *Coxiella burnetii*: Data from the United States National Health and Nutrition Examination Survey.**

**Charles F Dillon, Gwendolyn R Dillon**

**Table 1. *Coxiella burnetii* Seropositivity by History of Work in 17 Ocupational Groups.**

| Ocupational Groups: | n | Serology Positive | Percent |
|---|---|---|---|
| Health Services Workers | 397 | 6 | 1.51 |
| Office Workers | 865 | 14 | 1.62 |
| Mechanics, Repairers | 251 | 6 | 2.39 |
| Education, Teachers | 276 | 7 | 2.54 |
| Sales Workers | 618 | 17 | 2.75 |
| Food Manufacturing, Sales, & Services Workers | 459 | 14 | 3.05 |
| Professional Specialties | 402 | 14 | 3.48 |
| Protective Services (Police,1st Responders) | 83 | 3 | 3.61 |
| Other Machine Operators, Assemblers, Inspectors | 453 | 17 | 3.75 |
| Laborers | 191 | 8 | 4.19 |
| Executives & Management | 543 | 23 | 4.24 |
| Cleaning, Building Services | 221 | 12 | 5.43 |
| Textile, Apparel, Furnishings Machine Operators | 223 | 14 | 6.28 |
| Construction Workers | 350 | 22 | 6.29 |
| Transportation Workers | 306 | 20 | 6.54 |
| Personal Services Workers | 145 | 10 | 6.90 |
| Agricultural, Farm & Forestry Workers | 187 | 27 | 14.44 |

The occupational groups are not mutually exclusive, a sample person's lifetime history of work in any of the listed occupational categories is presented. Only crude percentages are presented as most ccupational group estimates were statistically unreliable. Positive *C. burnetii* serology is a Phase 1 or Phase 2 IgG IFA titer ≥ 1:16.

`

### Table 2. US Bureau of Labor Statistics 2003 Labor Force Participation Rates.

HOUSEHOLD DATA
ANNUAL AVERAGES

3. Employment status of the civilian noninstitutional population by age groups.
(Numbers in thousands)

Notes: Table numeric column 3 "Percent of Population" shows the US Labor Force Participation Rate percentages.The percentages are calculated by dividing numeric column 2 ("Total", the number of persons currently employed or actively looking for work) by column 1 (the total US "Civilian noninstitutional population"). [Numeric columns 1,4 & 5 are used to estimate the Employment to Population Ratio: only the percentage of those currently working-not used in study].

| | | 2003 | | | | | | |
|---|---|---|---|---|---|---|---|---|
| | | Civilian labor force | | | | | | |
| Age Groups | Civilian noninstitutional population | Total | Percent of population | Employed | | Unemployed | | Not in labor force |
| | | | | Total | Percent of population | Number | Percent of labor force | |
| TOTAL | | | | | | | | |
| 16 years and over | 221,168 | 146,510 | 66.2 | 137,736 | 62.3 | 8,774 | 6.0 | 74,658 |
| 16 to 19 years | 16,096 | 7,170 | 44.5 | 5,919 | 36.8 | 1,251 | 17.5 | 8,926 |
| 16 to 17 years | 8,561 | 2,857 | 33.4 | 2,312 | 27.0 | 545 | 19.1 | 5,704 |
| 18 to 19 years | 7,535 | 4,313 | 57.2 | 3,607 | 47.9 | 706 | 16.4 | 3,222 |
| 20 to 24 years | 19,801 | 14,928 | 75.4 | 13,433 | 67.8 | 1,495 | 10.0 | 4,874 |
| 25 to 54 years | 123,289 | 102,309 | 83.0 | 97,178 | 78.8 | 5,131 | 5.0 | 20,980 |
| 25 to 34 years | 39,021 | 32,343 | 82.9 | 30,383 | 77.9 | 1,960 | 6.1 | 6,678 |
| 25 to 29 years | 18,625 | 15,357 | 82.5 | 14,339 | 77.0 | 1,018 | 6.6 | 3,267 |
| 30 to 34 years | 20,396 | 16,986 | 83.3 | 16,044 | 78.7 | 941 | 5.5 | 3,411 |
| 35 to 44 years | 43,746 | 36,695 | 83.9 | 34,881 | 79.7 | 1,815 | 4.9 | 7,051 |
| 35 to 39 years | 21,050 | 17,571 | 83.5 | 16,663 | 79.2 | 908 | 5.2 | 3,480 |
| 40 to 44 years | 22,696 | 19,125 | 84.3 | 18,218 | 80.3 | 907 | 4.7 | 3,571 |
| 45 to 54 years | 40,522 | 33,270 | 82.1 | 31,914 | 78.8 | 1,356 | 4.1 | 7,252 |
| 45 to 49 years | 21,581 | 18,081 | 83.8 | 17,325 | 80.3 | 756 | 4.2 | 3,500 |
| 50 to 54 years | 18,941 | 15,189 | 80.2 | 14,589 | 77.0 | 601 | 4.0 | 3,751 |
| 55 to 64 years | 27,728 | 17,312 | 62.4 | 16,598 | 59.9 | 713 | 4.1 | 10,416 |
| 55 to 59 years | 15,625 | 11,142 | 71.3 | 10,685 | 68.4 | 457 | 4.1 | 4,483 |
| 60 to 64 years | 12,103 | 6,170 | 51.0 | 5,913 | 48.9 | 257 | 4.2 | 5,933 |
| 65 years and over | 34,253 | 4,792 | 14.0 | 4,608 | 13.5 | 183 | 3.8 | 29,462 |
| 65 to 69 years | 9,591 | 2,627 | 27.4 | 2,515 | 26.2 | 112 | 4.2 | 6,964 |
| 70 to 74 years | 8,456 | 1,231 | 14.6 | 1,189 | 14.1 | 43 | 3.5 | 7,225 |
| 75 years and over | 16,207 | 934 | 5.8 | 904 | 5.6 | 29 | 3.1 | 15,273 |

**Table 3. US Bureau of Labor Statistics 2004 Labor Force Participation Rates.**

HOUSEHOLD DATA
ANNUAL AVERAGES

3. Employment status of the civilian noninstitutional population by age groups.
(Numbers in thousands)

Notes: Table numeric column 3 "Percent of Population" shows the US Labor Force Participation Rate percentages.The percentages are calculated by dividing numeric column 2 ("Total", the number of persons currently employed or actively looking for work) by column 1 (the total US "Civilian noninstitutional population"). [Numeric columns 1,4 & 5 are used to estimate the Employment to Population Ratio: only the percentage of those currently working-not used in study].

2004

| Age Groups | Civilian noninsti- tutional population | Civilian labor force | | Employed | | Unemployed | | Not in labor force |
|---|---|---|---|---|---|---|---|---|
| | | Total | Percent of population | Total | Percent of population | Number | Percent of labor force | |
| TOTAL | | | | | | | | |
| 16 years and over | 223,357 | 147,401 | 66.0 | 139,252 | 62.3 | 8,149 | 5.5 | 75,956 |
| 16 to 19 years | 16,222 | 7,114 | 43.9 | 5,907 | 36.4 | 1,208 | 17.0 | 9,108 |
| 16 to 17 years | 8,574 | 2,747 | 32.0 | 2,193 | 25.6 | 554 | 20.2 | 5,827 |
| 18 to 19 years | 7,648 | 4,367 | 57.1 | 3,714 | 48.6 | 653 | 15.0 | 3,281 |
| 20 to 24 years | 20,197 | 15,154 | 75.0 | 13,723 | 67.9 | 1,431 | 9.4 | 5,043 |
| 25 to 54 years | 123,410 | 102,122 | 82.8 | 97,472 | 79.0 | 4,650 | 4.6 | 21,288 |
| 25 to 34 years | 38,939 | 32,207 | 82.7 | 30,423 | 78.1 | 1,784 | 5.5 | 6,732 |
| 25 to 29 years | 18,985 | 15,569 | 82.0 | 14,615 | 77.0 | 955 | 6.1 | 3,415 |
| 30 to 34 years | 19,954 | 16,638 | 83.4 | 15,808 | 79.2 | 829 | 5.0 | 3,317 |
| 35 to 44 years | 43,226 | 36,158 | 83.6 | 34,580 | 80.0 | 1,578 | 4.4 | 7,068 |
| 35 to 39 years | 20,573 | 17,169 | 83.5 | 16,370 | 79.6 | 799 | 4.7 | 3,404 |
| 40 to 44 years | 22,653 | 18,989 | 83.8 | 18,210 | 80.4 | 779 | 4.1 | 3,664 |
| 45 to 54 years | 41,245 | 33,758 | 81.8 | 32,469 | 78.7 | 1,288 | 3.8 | 7,488 |
| 45 to 49 years | 21,886 | 18,310 | 83.7 | 17,586 | 80.4 | 724 | 4.0 | 3,577 |
| 50 to 54 years | 19,359 | 15,448 | 79.8 | 14,883 | 76.9 | 565 | 3.7 | 3,911 |
| 55 to 64 years | 28,919 | 18,013 | 62.3 | 17,331 | 59.9 | 682 | 3.8 | 10,906 |
| 55 to 59 years | 16,327 | 11,603 | 71.1 | 11,166 | 68.4 | 437 | 3.8 | 4,724 |
| 60 to 64 years | 12,592 | 6,410 | 50.9 | 6,166 | 49.0 | 245 | 3.8 | 6,182 |
| 65 years and over | 34,609 | 4,998 | 14.4 | 4,819 | 13.9 | 179 | 3.6 | 29,611 |
| 65 to 69 years | 9,800 | 2,710 | 27.7 | 2,614 | 26.7 | 96 | 3.5 | 7,090 |
| 70 to 74 years | 8,381 | 1,280 | 15.3 | 1,234 | 14.7 | 46 | 3.6 | 7,100 |
| 75 years and over | 16,429 | 1,007 | 6.1 | 971 | 5.9 | 36 | 3.6 | 15,421 |

**Table 4. Non-Employed Persons-Total Person-Years Worked.**

| Reason Not in Labor Force | n | Person-Years Worked |
|---|---|---|
| **Total:** | 1,930 | 33,072 |
| Caring for Home, Family | 297 | 1,717 |
| Going to School | 62 | 235 |
| Retired | 1,020 | 24,022 |
| Ill Health | 174 | 2,369 |
| On layoff | 41 | 362 |
| Disabled | 230 | 3,400 |
| Other Reasons | 106 | 967 |

1 person-year= 1 year an individual had worked.
